# Effectiveness, cost effectiveness and experiences of switching from intravenous to oral antibiotics in neonates with probable early onset sepsis: a systematic review

**DOI:** 10.1101/2025.10.20.25338361

**Authors:** Rebecca Whear, Rebecca Abbott, Harriet Aughey, Morwenna Rogers, Alison Bethel, Stuart Logan, Kelly Boxall, Jo Thompson Coon

## Abstract

**Objective:** To assess the effectiveness, cost-effectiveness and experiences of switching from intravenous antibiotics to oral antibiotics in neonates with suspected early onset sepsis (EOS).

**Design:** Systematic review.

**Patients:** Clinically stable term and later preterm neonates with suspected EOS.

**Interventions:** Oral antibiotics.

**Main outcome measures:** Mortality and morbidity, cost and resource use, family related and process outcomes and experiences of families and health care professionals.

**Information sources:** We searched Medline, Embase, PsycINFO, HMIC, SPP, CENTRAL, CINAHL and Conference Abstracts (via Web of Science) in April 2025. Database searches were supplemented with citation chasing, contacting authors and website searches.

**Methods:** Screening, data extraction and critical appraisal was performed in duplicate. Due to heterogeneity of outcomes, timepoints and lack of reported data relating to the data distribution, a narrative synthesis was performed. The review was informed by public collaborators interested in maternal health.

**Results:** After de-duplication, 3803 titles and abstracts were screened for relevance with 27 assessed for eligibility at full text. Following title and abstract screening, one randomised controlled trial (504 neonates) and three observational studies (795 neonates) were included. Length of stay was reduced by between 1 to 4 days across all four studies. No differences were observed between study arms in those reporting on readmission rates, reinfection or mortality.

**Conclusions:** This systematic review suggests that switching clinically stable term and late pre-term neonates with suspected EOS from intravenous to oral antibiotics can reduce hospital stay and healthcare costs without increasing morbidity, mortality, or readmission.

**Key messages:**

- What is already known on this topic

Intravenous antibiotics are commonly prescribed in neonatal care to treat suspected early onset sepsis (EOS). Distinguishing infectious from non-infectious cases is difficult, often leading to unnecessary intravenous antibiotic treatment in uninfected neonates. A recent systematic review of oral antibiotics for neonatal infections concluded that in general, adequate serum levels can be achieved after oral administration of antibiotics in neonates.

- What this study adds

There is promising evidence that switching clinically well, term and late preterm neonates with suspected EOS from intravenous to oral antibiotics reduces duration of hospital stay with no impact on readmissions, clinical deterioration, mortality, or reinfection.

- How this study might afect research, practice or policy

Future research should focus on longer-term outcomes such as effects on the gut microbiome and later childhood health, evaluations of cost-effectiveness, incorporating both healthcare system and family perspectives, and ensure that equity considerations are fully addressed.

## Introduction

Antibiotics are among the most prescribed medications in neonatal care, with many neonates receiving them due to suspected bacterial infections during their first few weeks of life.^1^ Of particular concern is *early onset sepsis* (EOS) which refers to suspected neonatal infections within the first 72 hours of life, which untreated can lead to catastrophic results^1^. The signs of infection are relatively non-specific and there is a lack of diagnostic tests which can reliably discriminate between babies with a bacterial infection and those without or in whom infection has been effectively treated, leading to both the unnecessary initiation and prolongation of antibiotic treatment.^2^ An EOS calculator^3^, the Kaiser Permanente Sepsis Risk Calculator (SRC) has been introduced in recent years to statistically stratify the risk of neonatal infection on a case-by-case basis, thereby helping to rationalise the initiation of antibiotic treatment. However, the use of this in clinical practice varies across the UK. Although the incidence of suspected infection is highest amongst premature infants, a substantial number of term and late premature infants are also affected. A recent study in London reported that of infants >=34 weeks gestation, 11.6% received antibiotics within the first 24 hours of life. There were large differences between units in the proportion of infants treated, with those basing decisions on the SRC reporting substantially lower rates than those using the NICE guidance (7.2% vs 14.8%)^3^.

For most infants, antibiotic therapy can be discontinued after 36–48 hours if they are clinically well and have serial low C-reactive protein (CRP) results, and negative blood cultures. In a small proportion (0.6% in the London study)^3^, infection is confirmed but in around 25-30%, corresponding to approximately 1.5% of all term and late preterm infants, the infants are clinically well but bacterial infection cannot be ruled out. Standard practice is to continue intravenous antibiotics for these neonates for 5–7-days.^4^ Switching from intravenous to oral antibiotics for these infants could offer several benefits, particularly the possibility of earlier hospital discharge which would be advantageous for both families and healthcare systems.^5^

A recent systematic review of oral antibiotics for neonatal infections concluded that in general, adequate serum levels can be achieved after oral administration of antibiotics in neonates.^6^ Furthermore, gentamicin, one of the antibiotics which is routinely used to treat neonates intravenously, carries a risk of hearing loss in those with genetic susceptibility.^7^ Switching to oral antibiotics may reduce these risks.

This review aimed to address the following research questions:

- What is the impact on morbidity and mortality, family outcomes and cost and resource use of switching from intravenous to oral antibiotics in neonates with suspected EOS?
- What are the views, experiences and perceptions of health care professionals, parents and families of switching from intravenous antibiotics to oral antibiotics in neonates with suspected EOS?

## Methods

The protocol was prospectively registered on PROSPERO CRD420251044158 and is reported following PRISMA guidelines.^8^

### Identification of studies

The search strategy was developed by an information specialist in collaboration with the project team. We searched Medline, Embase, PsycINFO, HMIC and SPP (via Ovid), CENTRAL (via the Cochrane Library) CINAHL (via EBSCOhost) and Conference Abstracts (via Web of Science) using terms for oral drug administration, antibiotics and neonates. The search strategy for Embase is shown in Box 1. We also searched the Cochrane Database of Systematic Reviews and Epistemonikos for related reviews, and clinical trials registries (clinicaltrials.gov and the WHO International Clinical Trials Registry Platform (ICTRP)) for ongoing trials and Google, using the advanced search, for reports and, evaluations in specific web domains including nhs.uk and gov.uk. We also searched the websites of The Neonatal Society, the British Association of Perinatal Medicine, the Royal College of Paediatrics and Child Health, the European Society of Paediatric and Neonatal Intensive Care, the Union of European Neonatal & Perinatal Societies, the European Society for Pediatric Research and the American Academy of Pediatrics. Searches were conducted in April 2025.

We carried out forwards and backwards citation searching using Web of Science and contacted the authors of included papers and conference abstracts in June 2025.

### Study selection

Title and abstract screening was performed independently by two reviewers (MR, AB, RW, RA) with disagreements resolved through discussion or referral to a third reviewer. The full texts of all potentially relevant citations were retrieved and the eligibility criteria applied using the same method. Studies were eligible for inclusion if they met the following criteria:

#### Population

Clinically stable, term and late pre-term (greater than 35 weeks gestation) babies with suspected EOS (within the first 72 hours of life).

#### Intervention

Switch to oral antibiotics.

#### Comparator

Remain on intravenous antibiotics.

#### Outcomes

Mortality and morbidity, cost and resource use, process outcomes, family-related outcomes and the views, perceptions and experiences of health care professionals, parents and families of switching from intravenous to oral administration of antibiotics. Full details can be found in the protocol^9^.

#### Setting

Hospitals.

#### Study design

Any comparative study design collecting relevant, empirical data including service evaluations and quality improvement projects. Qualitative studies on the views, perceptions and experiences of switching to oral antibiotics. Conference abstracts were eligible for inclusion if sufficient information was provided. Protocols and ongoing trials from clinical trial registries were also eligible for inclusion.

We applied no date, geographical or language restrictions.

Endnote 21 software was used to support record management and study selection.

### Data extraction and quality appraisal

Data on sample characteristics, intervention and comparator details, reported outcomes and equity characteristics were extracted by one reviewer (RW) and checked by a second (JTC, RA). For qualitative studies, we also planned to collect data on the amount of available data relevant to the research questions available and the themes or ideas presented^9^.

Quantitative studies were appraised using the Effective Public Health Practice Project (EPHPP) tool^10^ by one reviewer (RW) and checked by a second (RA) with disagreements resolved through discussion. Authors of included studies were contacted for additional data (mean, standard deviations for duration of hospital stay); all provided the relevant data. We requested additional data for missing outcomes (parental satisfaction, gut microbiome and cost effectiveness) from one study^11^; these were not available.

### Data analysis and presentation

Due to a lack of events (for categorical outcomes), a large variation in time points for other variables of interest (such as mortality), and/or a lack of information reported regarding the distribution of the outcome variable, we could not statistically pool the data. We therefore conducted a narrative synthesis to interpret the findings which is reported according to SWiM guidelines.^12^

We structured the synthesis according to the four outcome categories. We tabulated the findings reporting the RCT first, followed by findings from the observational studies, noting similarities and differences where appropriate.

The absence of eligible qualitative studies precluded any qualitative or mixed methods synthesis.

### Health inequalities

We used the PROGRESS-Plus framework^13^ to consider variations across populations and subgroups.

### Patient and public involvement

We convened a group of public collaborators interested in maternal health who informed the development of the protocol, contributed to the interpretation of findings and helped to identify implications for research and practice from the patient perspective.

## Results

A total of 5528 records were retrieved from the databases and searches of the clinical trials registries (Figure 1). No further studies were retrieved through website searches or citation chasing. After duplicate records were removed, 3803 titles and abstracts were screened initially with another 139 screened after citation searching. Twenty-seven full text documents were retrieved and assessed for eligibility. Following the screening process, four studies were included. Reasons for exclusion at full text are shown in the PRISMA flow diagram (Figure 1); excluded studies are provided in Supplementary Table 1.

**Figure 1:**
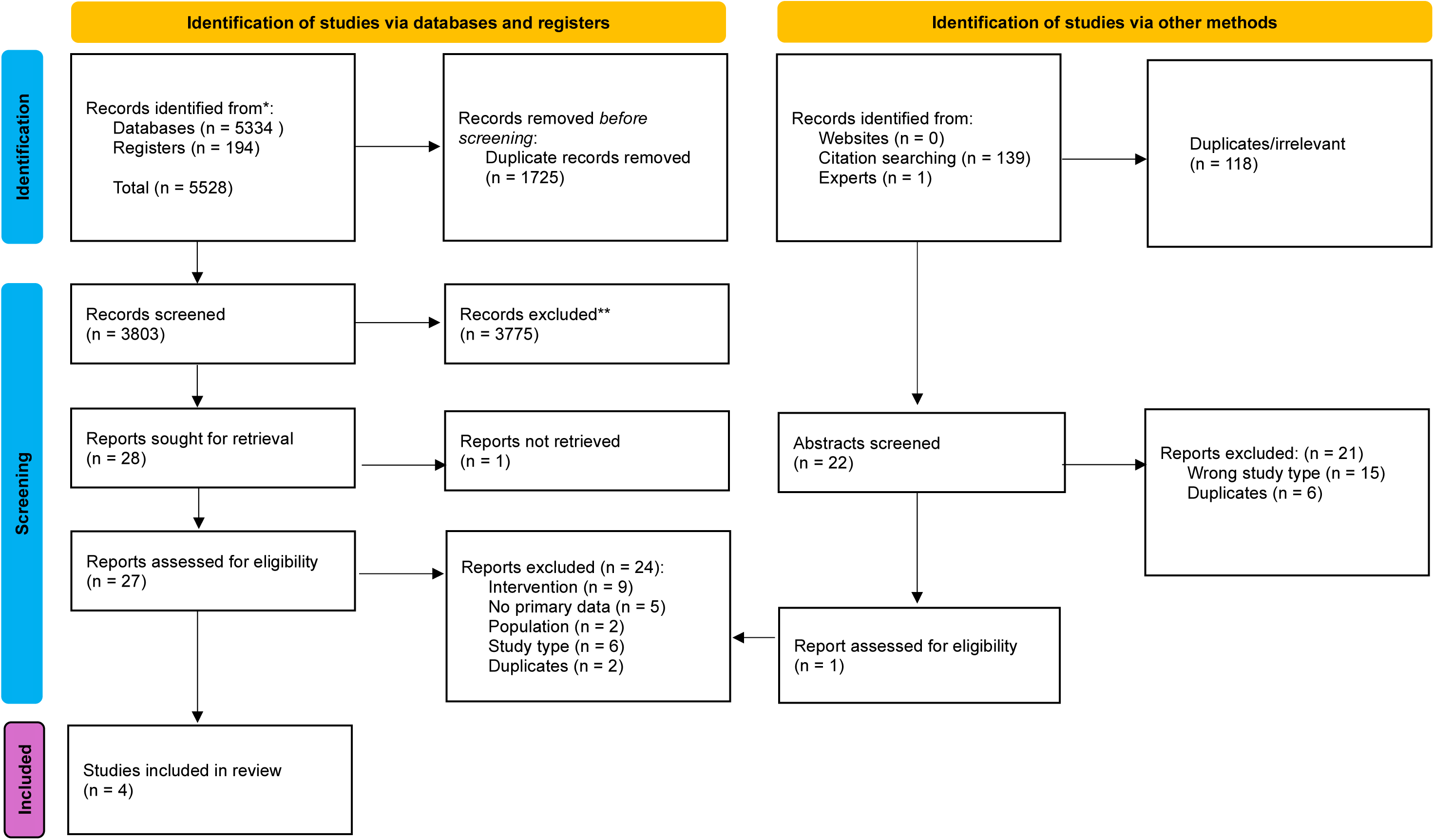
PRISMA flow chart

### Characteristics of included studies

Characteristics of the four included studies are shown in Table 1. One randomised controlled trial (RCT) conducted in the Netherlands^11^, and three observational studies from Sweden^14^, Denmark^15^ and Italy^16^. In total the studies included 1209 neonates: 504 in the RCT^11^ and 705 in the three observational studies^14–16^. Two studies assessed the impact of new antibiotic guidance (which involved a switch to oral) for suspected EOS in neonates following implementation in their localities by comparing data from the new approach to retrospective records.^14^ ^15^ The principal focus of the study by Gyllensvard and colleagues^14^ was to evaluate whether C-reactive protein could be a useful guide to help clinicians to discontinue antibiotic therapy, and the switch to oral antibiotics was an extra precaution. Neonates were discharged home once they had been shown to tolerate oral administration and had no other reason to be hospitalised in two of the studies.^11^ ^15^ Three studies described the safety protocol used to follow up neonates in either both arms^11^ ^14^ or the intervention arm only^15^ (Table 1).

**Table 1:**
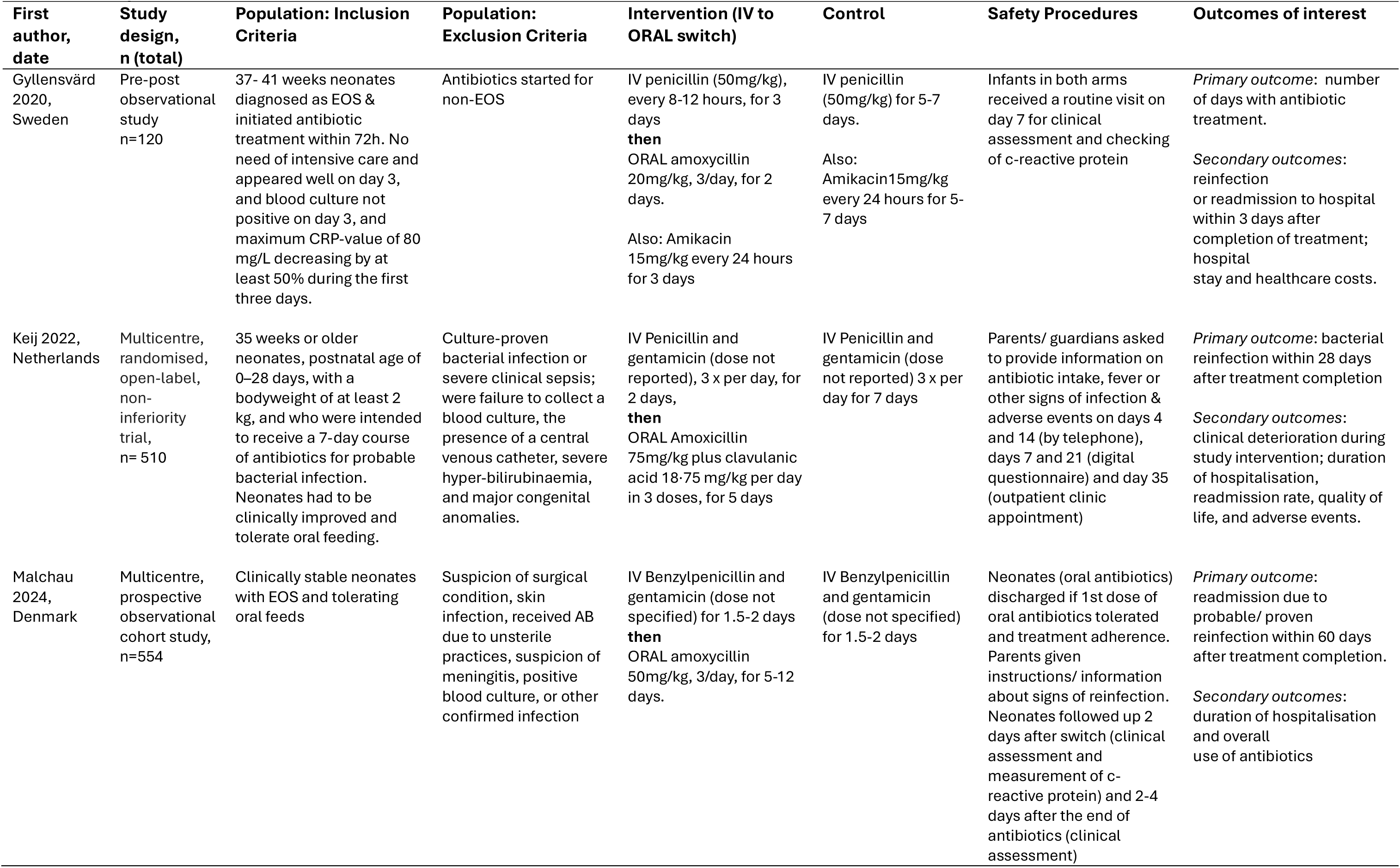
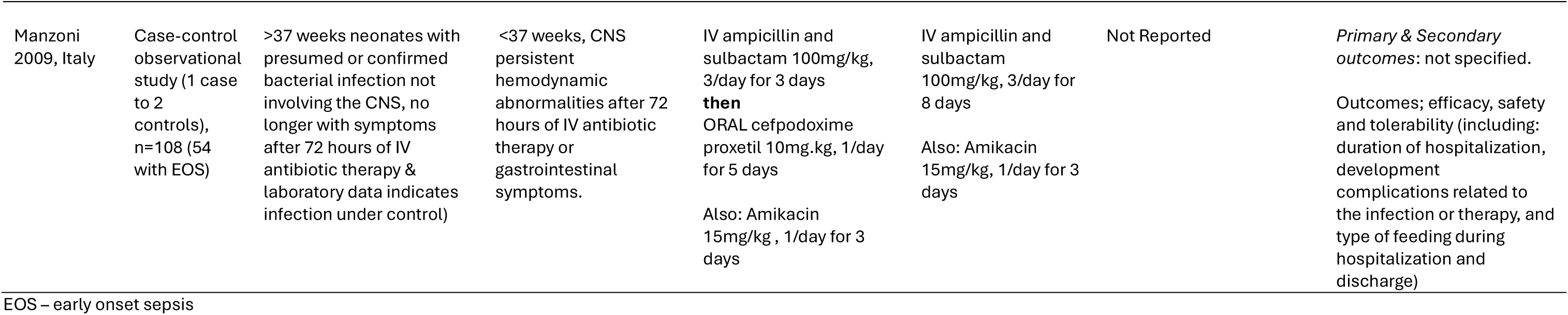
Study characteristics.

Narrow, well-defined eligibility criteria meant that the recruited neonates were similar across the studies in terms of gestation, body weight and gender (Table 2). Maternal risk factors for neonatal EOS, where reported^11^ ^14^ ^16^, were also similar between study arms and across studies (Table 3, presented according to risk factors identified within National Institute for Health and Care Excellence (NICE) Guideline NG195^17^). Equity characteristics were not well reported. Studies provide information on age and gender, but other equity characteristics were not reported, for example, ethnicity, socioeconomic status, place of residence (i.e. location, urban/rural, type of housing), culture, religion or social networks.

**Table 2:** Sample characteristics.

| Study | Study Design |  | Sample size | Gestational age in weeks<br>Median (IQR, unless specified) | Birthweight (g)<br>Mean (SD) | Gender (% Female) | Ethnicity | Delivery method | Study Quality |
| --- | --- | --- | --- | --- | --- | --- | --- | --- | --- |
| Keij 2022 | RCT | *Intervention* | 252 | 40.3 (IQR 39.3–41.0) | 3592 (482) | 41 | NR | Vaginal 159 (63%),<br>Instrumental 35 (14%),<br>Caesarean 58 (23%) | Moderate |
|  |  | *Control* | 252 | 40.4 (IQR 39.3–41.0) | 3643 (561) | 42 |  | Vaginal 157 (62%),<br>Instrumental 28 (11%),<br>Caesarean 67 (27%) |  |
| Gyllensvärd 2020 | Pre-post observational study | *Intervention* | 59 | 40 (IQR 39-41) | 3899 (497) | 46 | NR | NR | Moderate |
|  |  | *Control* | 61 | 40 (IQR 39-40) | 3768 (559) | 43 | NR | NR |  |
| Malchau 2024 | Multicentre, prospective observational cohort study | *Intervention* | 478 | 40+3 (IQR 40+1- 41+5) | 3789 (500) | 38 | NR | NR | Moderate |
|  |  | *Control* | 53 | 40+3 (IQR 39+1- 41+5) | 3643 (519) | 31 | NR | NR |  |
| Manzoni 2009 | Case-control observational study | *Intervention* | 17 | 39.6 (+-2) | 3055 (+-590) | 50 | 94% caucasian | 78% vaginal | Weak |
|  |  | *Control* | 37 | 39.2 (+-2) | 3100 (+-545) | 50 | 94% Caucasian | 73% vaginal |  |

**Table 3:**
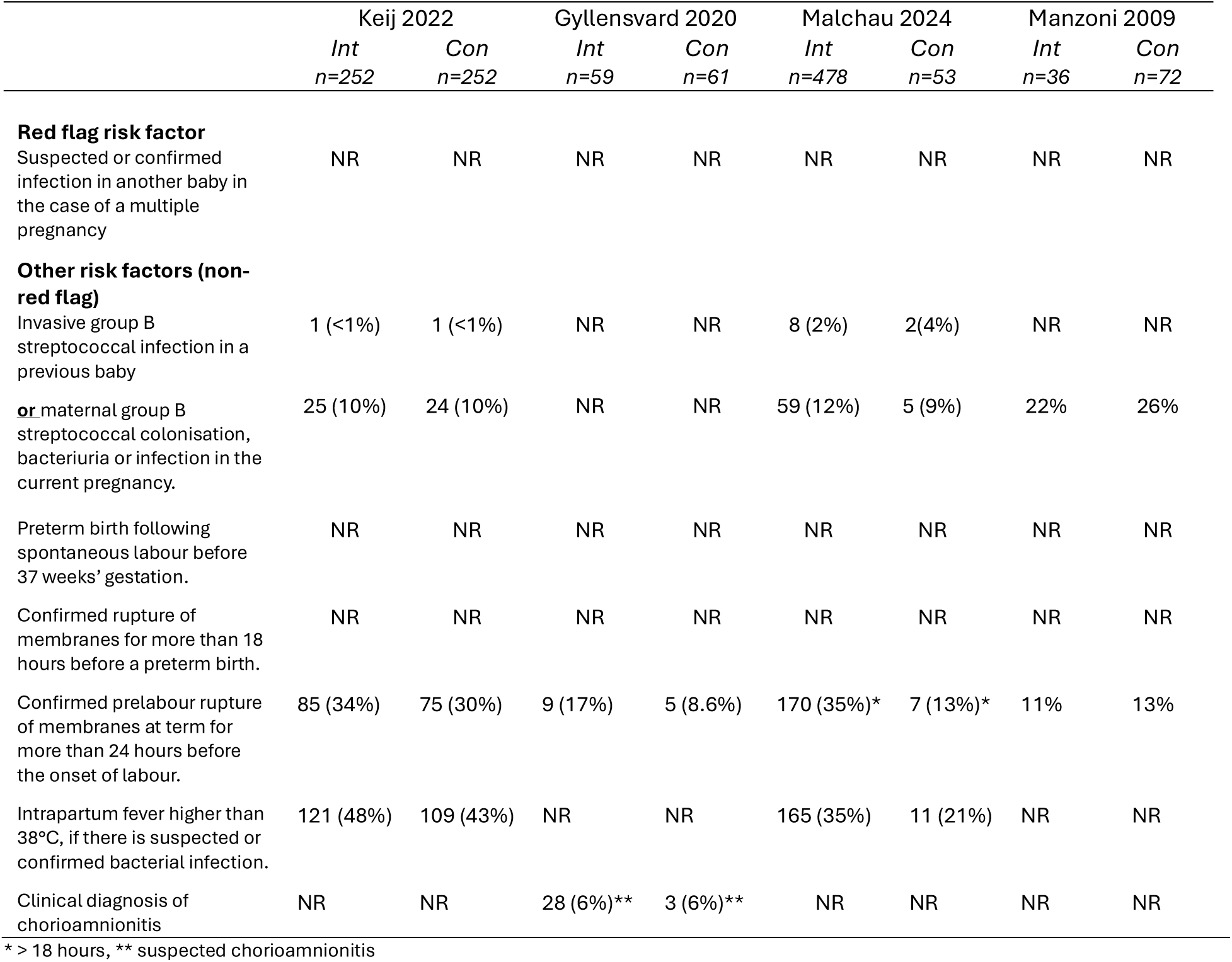
Maternal characteristics presented according to NICE Guideline No. 195.

### Quality of included studies

We rated the RCT^11^ and two of the observational studies^14^ ^15^ as moderate quality and the other observational study as weak.^16^ The individual breakdown by study quality components is shown in Table 4. All studies were rated as ‘weak’ with regards to participants and/or assessors being blinded to the intervention, which prevented any study from being rated overall as strong. Withdrawals and/or dropouts were not reported in the study by Manzoni and colleagues^16^.

**Table 4:**
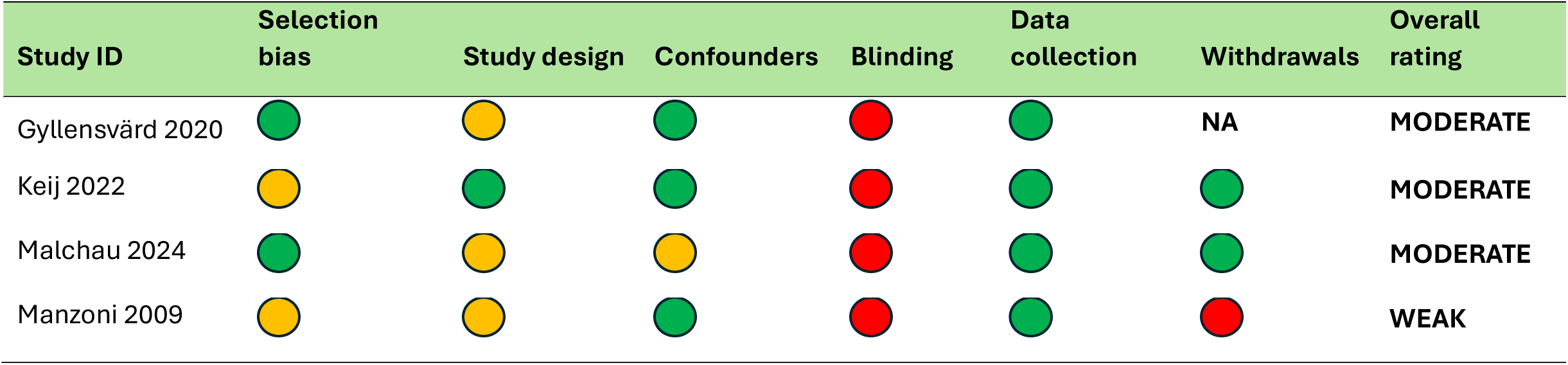
Quality appraisal summary for included studies.

## Synthesis by outcome

### Mortality and morbidity

Mortality and morbidity data are presented in Table 5.

**Table 5:**
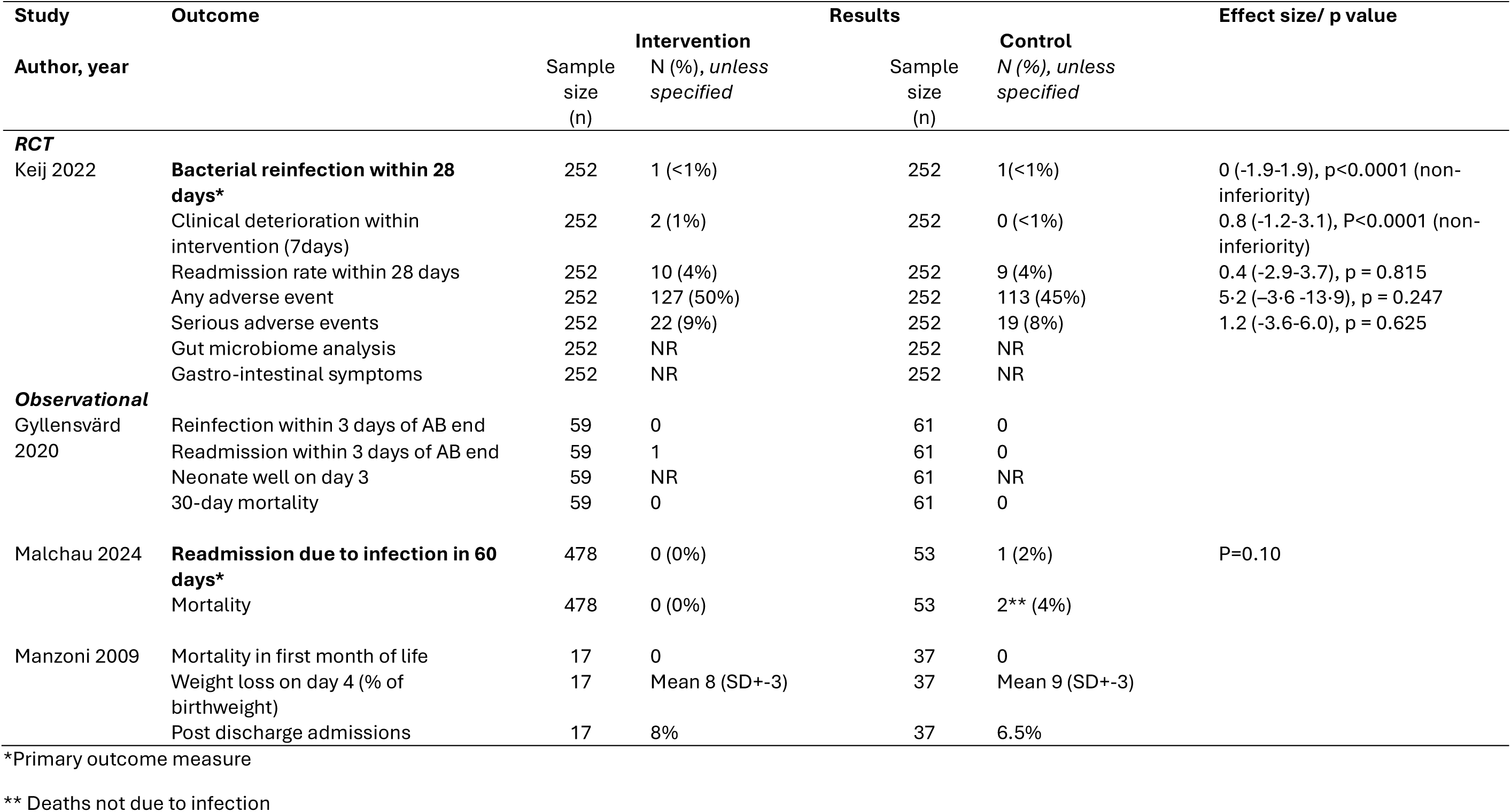
Mortality and morbidity outcomes.

The primary outcome measure in the RCT was bacterial reinfection at 28 days. No difference was found between study arms with only 1 neonate in each arm experiencing re-infection by this timepoint^11^. One observational study reported reinfection rates, albeit at an earlier timepoint (3 days post treatment) and found no difference between arms (no neonates in either arm).^14^

Readmission rates were measured at different time points in each study. In the RCT, readmission within one month of treatment was 4% in both intervention and control RCT (0.4 (-2.9 to 3.7), p = 0.815).^11^ The observational studies reported similar or lower readmission rates at 3 days post treatment^14^, within 60 days of treatment^14^, and at an unspecified time point^16^; there were no statistical differences between arms reported in any study.

The RCT also reported on clinical deterioration between arms (0.8 (-1.2 to 3.1), p<0.0001 (non-inferiority analysis), any adverse event (5·2 (–3·6 to 13·9), p = 0.247) and serious adverse events (1.2 (-3.6 to 6.0), p = 0.625)^11^; these outcomes were not measured in any of the observational studies. One observational study^16^ reported on weight loss at day 4 post treatment, with 8% loss reported in both arms (statistical data not provided).

Mortality rates at various time points (at 30 days^14^, in the first month of life^16^, and at an unspecified time^15^) were reported in the three observational studies. Deaths were reported in one study^15^; two neonates died (not from infection) in the IV treatment arm.

### Cost and resource use

Cost and resource use data are presented in Table 6 and were measured in a variety of ways across the studies including length of hospitalisation, contact with health professionals, and duration of antibiotic treatment.

**Table 6:** Cost and resource use.

| Study | Outcome | Intervention |  | Results |  | Effect size/ p value |
| --- | --- | --- | --- | --- | --- | --- |
| Author, year |  | Sample size<br>(n) | Days (Median,<br>IQR) or cost,<br>unless specified | Sample size<br>(n) | Control<br>Days (n), unless<br>specified |  |
| **RCT** |  |  |  |  |  |  |
| Keij 2022 | Duration of hospital stay | 252 | 3.4 (3.0-4.1) | 252 | 6.8 (6.5-7.0) | P<0.0001 |
|  | Proportion receiving health<br>care professional visit<br>between day 7-35 | 252 | 73 (29%) | 252 | 58 (23%) | 6.0 (-1.7-13.6), p = 0.128 |
|  | Cost effectiveness | 252 | NR | 252 | NR |  |
| **Observational** |  |  |  |  |  |  |
| Gyllensvärd<br>2020 | **Duration of antibiotic**<br>**treatment (days)\*** | 59 | Median 5 (IQR 5-6) | 61 | Median 7 (IQR 7-8) | P<0.001 |
|  | Cost saved per day/patient | 59 | 1350 Euros | 61 |  |  |
|  | Cost saved per annum | 59 | 122000 Euros | 61 |  |  |
|  | Days saved per patient | 59 | 2 | 61 | 0 |  |
|  | Number of days IV AB | 59 | Median 3 (IQR 3-4) | 61 | Median 5 (IQR 5-7) | P<0.001 |
|  | Number of days Oral AB | 59 | Median 2 (IQR 2-3) | 61 | Median 0 (IQR 0-4) | P=0.064 |
|  | Duration of hospital stay | 59 | 5 (4-6) | 61 | 7 (6-8) | P<0.001 |
| Malchau 2024 | Duration of hospital stay | 478 | 3 (2.5-3.5) | 53 | 7.2 (7.0-7.5) | P<0.001 |
|  | Duration of treatment | 478 | Median 7 (IQR 7-7) | 53 | Median 7 (IQR 7-7) | P=0.19 |
| Manzoni 2009 | Duration of hospital stay | 17 | Mean 7 (SD+-4) | 37 | Mean 8 (SD+-4) |  |
|  | Number of IV injections | 17 | Mean 9 (SD+-5) | 37 | Mean 14.9 (SD+-9) |  |

\*Primary outcome measure

As might be expected with an intervention that encourages hospital discharge for neonates randomised to switch to oral antibiotics, the RCT showed that duration of hospital stay was significantly shorter for those neonates in the intervention (4.12 days) arm compared to control (6.71 days) a mean difference of 2.59 days less for those on oral antibiotics (CI-2.93 to-2.25).^11^ Reductions of a similar magnitude were reported in two of the three observational studies (Figure 2 and Table 7).^14^ ^15^ Possibly as a consequence of a difference in the discharge strategy used, Malchau and colleagues report a greater reduction in the mean number of days in hospital for the intervention group.^15^ In this study, infants were switched to oral antibiotics *within* 36-48 hours after the start of IV antibiotics. Whereas in the other three studies infants in the intervention group were only switched to oral antibiotics *after* two-three days (48-72hours) of IV antibiotics according to their local policies.

**Figure 2:**
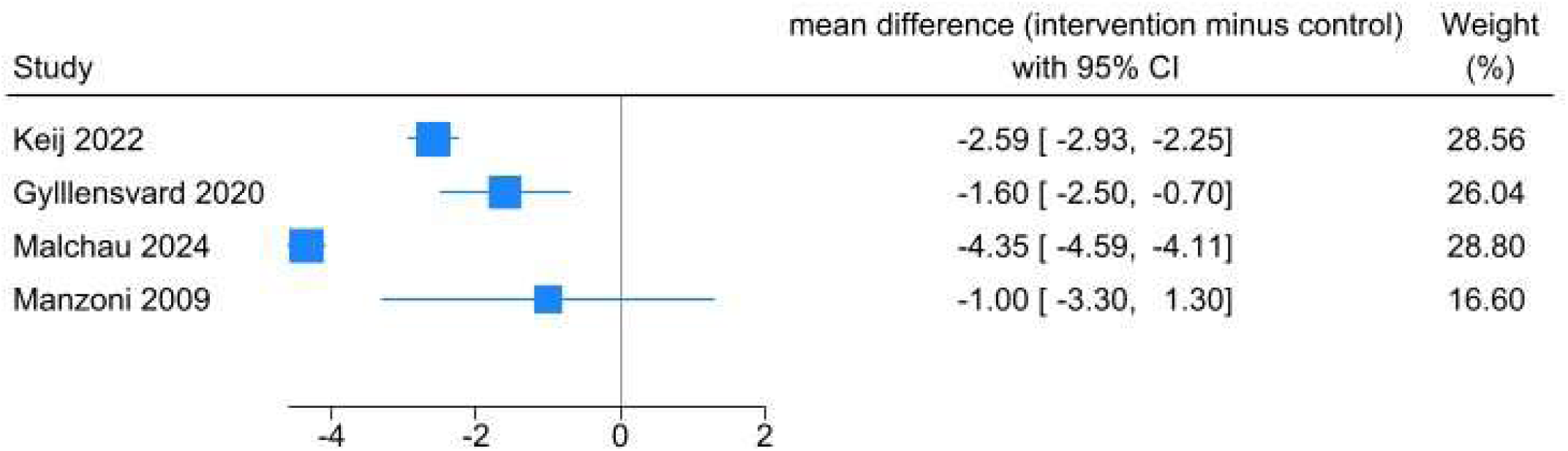
**Forest plot of effectiveness on mean days of stay in hospital**

**Table 7:** Mean length of stay.

| **Study** | **Design** | **Intervention**<br>**n** | **LOS**<br>**mean (SD)**<br>**days** | **Control**<br>**n** | **LOS**<br>**mean (SD)**<br>**days** | **Mean diff** | **95% CI** |
| --- | --- | --- | --- | --- | --- | --- | --- |
| Keij 2022 | RCT | 253 | 4.12 (2.33) | 252 | 6.71 (1.47) | -2.59 | -2.93, -2.25 |
| Gyllensvard 2020 | Cohort | 59 | 5.63 (2.86) | 61 | 7.23 (2.13) | -1.6 | -2.51, -0.69 |
| Malchau 2024 | Cohort | 478 | 3.15 (0.76) | 53 | 7.5 (1.41) | -4.35 | -4.59, -4.11 |
| Manzoni 2009 | Case control | 17 | 7 (4) | 37 | 8 (4) | -1 | -3.35, 1.35 |

LOS – length of stay

Contact with health professionals in the first 35 days post treatment was slightly higher in the intervention arm compared to control in the RCT (29% v 23%) but the difference was not statistically significant (6.0 (-1.7-13.6), p = 0.128).^11^

Two of the observational studies provided information on duration of antibiotic treatment. In one, the median duration of antibiotic treatment was significantly lower in the intervention arm (5 days (IQR 5 to 6) than the control arm (7 days (IQR 7 to 8) (p<0.001).^14^ In the other, there was no significant difference between the duration of antibiotic use between arms.^16^

Using the data on duration of hospital stay, Gyllensvard and colleagues^14^, calculated that the cost saved per day per patient was Eu1350 for those who switched to oral antibiotics, amounting to Eu122,000 saved per annum in their hospital. The cost-effectiveness outcome data collected within the RCT^11^ have not yet been published.

### Process outcomes

Process outcomes were reported in three studies and are presented in Table 8.

**Table 8:** Process outcomes.

| Study | Outcome | Results |  |  |  | Effect size/ p value |
| --- | --- | --- | --- | --- | --- | --- |
|  |  | Intervention |  | Control |  |  |
|  |  | Sample size | N (%), unless specified | Sample size | N (%), unless specified |  |
| **RCT** |  |  |  |  |  |  |
| Keij 2022 | Number of cannula reinsertion attempts during treatment | 252 | NA | 252 | Median 2 (IQR 1-3) |  |
|  | Serious adverse events | 252 | 22 (9%) | 252 | 19 (8%) | 1.2 (-3.6-6.0), p = 0.625 |
|  | Protocol violation | 252 | 18 (7%) | 252 | 53 (21%) | -13.9 (-20—7.8), p <0.0001 |
| **Observational** |  |  |  |  |  |  |
| Malchau 2024 | Treatment discontinuation | 478 | 0 | 53 | 0 |  |
|  | Oral medication accepted by neonate | 478 | 478 (100%) | 53 | NA |  |
|  | * Full course of antibiotics for suspected EOI (regional data) | 26114 | 309 (1.2%) | 26445 | 328 (1.2%) |  |
| Manzoni 2009 | Complications of the treatment | 17 | 0 | 37 | 0 |  |

EOI – early onset infection; \* Proportion of term-born neonates in region who received a full course of antibiotics, started before 7 days of life, due to EOI (method of AB usage not validated)

Significantly lower rates of protocol violations were reported in the intervention arm of the RCT compared to the control arm; 7% vs 21% (-13.9 (-20—7.8), p <0.0001).^11^ In the control arm, the majority of these related to non-completion of the full antibiotic course as intended. Treatment of 49 neonates in the control arm was not completed because of difficulty in venous access, and for 33 (67%) of these, treatment was continued orally. Relatedly, the authors reported that neonates in the control had a median of 2 (IQR 1-3) cannula reinsertion attempts (this outcome was not applicable in the intervention arm).^11^

Similarly, the observational studies reported no difference in treatment discontinuation^15^ or complications^16^ with both reporting zero instances across both arms for these two outcomes.

### Family-related

Family related outcomes are presented in Table 9 and were reported in two studies. No statistically significant differences were observed in the parental reported sleep quality of the neonates at 1 week (83% vs 78% reporting good sleep quality (5.6 (-1.4 to12.5), p = 0.115) or 1 month (71% vs 67% reporting good sleep quality (6.0 (-2.2 to14.1), p = 0.150) between study arms in the RCT.^11^ There were also no reported difference in the proportion of neonates exclusively breastfeeding at 1 month between intervention and control, 47% v 46% (0.4 (-8.3-9.1), p = 0.929).^11^ Manzoni and colleagues reported higher rates for exclusive breastfeeding at discharge for those in the intervention arm (94%, n=17) compared to control (76%, n=37), but no statistical data were provided.^16^

**Table 9:** Family-related.

| Study | Outcome | Results |  |  |  | Effect size/ p value |
| --- | --- | --- | --- | --- | --- | --- |
| Author, year |  | Intervention |  | Control |  |  |
|  |  | Sample size<br>(n) | N (%), *unless specified* | Sample size<br>(n) | N (%), *unless specified* |  |
| **RCT** |  |  |  |  |  |  |
| Keij 2022 | Exclusively breastfed at 1 month | 252 | 118 (47%) | 252 | 117 (46%) | 0.4 (-8.3-9.1), p = 0.929 |
|  | Sleep quality good at 1 week | 252 | 210 (83%) | 252 | 196 (78%) | 5.6 (-1.4-12.5), p = 0.115 |
|  | Sleep quality good at 1 month | 252 | 180 (71%) | 252 | 165 (66%) | 6.0 (-2.2-14.1), p = 0.150 |
|  | Parental satisfaction | 252 | NR | 252 | NR |  |
| **Observational** |  |  |  |  |  |  |
| Manzoni, 2009 | Type of feeding in hospital | 17 | NR | 37 | NR |  |
|  | Exclusively breastfed at discharge | 17 | 94% | 37 | 76% |  |

### Experiences

None of the included studies reported on the experience of switching to oral antibiotics from family or clinician perspectives

### Patient and public involvement

During development of the protocol, the public collaborators were interested to understand a) the safeguards that might be put in place to ensure that barriers to accessing help after the baby had gone home were minimised; b) whether families were given a choice and that consideration was given as to whether home was the safest/most practical place for them; and c) whether families preferences were listened to. They also suggested several outcomes of interest e.g. long-term impact on gut biome, parents’ perspectives on antibiotic use, costs to the family and 111 and emergency department visits. On sharing the results and discussion points, the group highlighted two areas for future research: parent/family experience of switching to oral antibiotics and the impact of antibiotics on breastfeeding and decisions around feeding, as well as emphasising the need for a clear focus on safety and safeguarding.

## Discussion

This is the first systematic review to synthesise the evidence on switching from intravenous antibiotics to oral antibiotics in clinically well, term (>37 weeks) and late preterm (>35 weeks) neonates with suspected EOS. The review included four studies: one randomised controlled trial^11^ (504 patients) and three observational studies^14^ ^16^ (795 patients). There is promising evidence that switching clinically well, term and late preterm neonates with suspected EOS from intravenous to oral antibiotics reduces duration of hospital stay with no impact on readmissions, mortality, or reinfection.

### Interpretation of findings

The RCT reported non-inferiority of oral antibiotic therapy compared with full-course IV therapy for reinfection at 28 days, with very low absolute event rates^11^. This finding was consistent with the three observational studies, which also reported no excess mortality or readmissions. Importantly, hospital stay was consistently reduced across all studies, with potential benefits for both the healthcare system and family wellbeing.

While adverse events and clinical deterioration were rare and did not differ between treatment groups, only the RCT systematically assessed these outcomes. Family-related outcomes including breastfeeding and infant sleep showed no differences between RCT arms, but data were not provided in the observational studies. Crucially, no included study reported qualitative data on parent or clinician experiences, despite these being important determinants of acceptability and implementation.

Cost savings were reported in two studies, consistent with reductions in length of hospitalisation, however, no full cost-effectiveness analyses were available.

### Strengths and weaknesses

The strength of this review lies in its robust methods as well as the input from patient and public contributors. However, the evidence base is limited. Only four studies were eligible for inclusion, and these varied in design and were rated as moderate or weak quality. Much of the evidence is drawn from a single RCT which, although relatively large, was open label in design. Furthermore, we lack data from the RCT on parental satisfaction, gut microbiome, gastrointestinal symptoms, and detailed cost-effectiveness.

Pooling of the results from the four studies was not possible due to heterogeneity of outcomes and timepoints and missing data relating to the data distribution. Family-related outcomes were minimally reported across the four studies. Follow-up periods were relatively short, and longer-term outcomes such as the impact on the gut microbiome or later childhood health were absent. Furthermore, all studies were conducted in European countries, limiting applicability to other healthcare contexts. Importantly, equity-related characteristics were poorly reported, with little or no data on ethnicity, socioeconomic status, or geography, raising concerns about applicability across diverse populations.

These studies were conducted in high-income European settings. Differences in population characteristics, maternal risk factors for infection, and the organisation of perinatal care are possible, and careful consideration is therefore needed when interpreting their relevance for UK practice.

### Comparison with the existing literature

These findings align with earlier pharmacokinetic studies^6^ suggesting that adequate serum concentrations of oral antibiotics can be achieved in neonates. However, it is worth noting that the neonatal literature on this topic remains sparse, with the absence of robust qualitative research on parental perspectives an important gap.

### Implications for practice and policy

Switching from intravenous to oral antibiotics represents a recognised antimicrobial stewardship strategy and is a current priority for the UK Health Security Agency.^18^ Evidence from broader populations has demonstrated multiple benefits of IV-to-oral switch, including lower risk of bloodstream and catheter-related infections, reductions in equipment use, costs and environmental impact, shorter hospital stays, improved comfort and mobility for patients, and more efficient use of nursing time. For clinicians and policymakers, the available evidence provides cautious support for IV-to-oral antibiotic switch strategies in stable term and late pre-term neonates, provided appropriate safety and governance processes are in place. Concerns have been raised ^14^ ^5^ that the easier administration of oral antibiotics could encourage greater overall use; any adoption of this approach should therefore be embedded within strong antimicrobial stewardship. Implementation also requires robust follow-up systems, attention to family needs and equitable access.

### Implications for research

The most pressing research priority is the development of better diagnostic tools to distinguish which neonates genuinely have bacterial infection, to avoid unnecessary antibiotic exposure in those who do not. Regarding a switch therapy for this cohort, future research should focus on longer-term outcomes such as effects on the gut microbiome and later childhood health. Evaluations of cost-effectiveness, incorporating both healthcare system and family perspectives, would provide important insights for policy. Finally, future research should explore parental and clinician experiences of early discharge and oral therapy, and ensure that equity considerations such as socioeconomic status, ethnicity, and access to follow-up care are fully addressed.

## Conclusions

This systematic review suggests that switching clinically stable term and late pre-term neonates with suspected EOS from intravenous to oral antibiotics can reduce hospital stay and healthcare costs without increasing morbidity, mortality, or readmission. UK studies^3^ suggest that this policy change would affect approximately 1.5% of all term and late preterm infants equating to 9,000 infants per year in the UK.

## Data Availability

All data produced in the present work are contained in the manuscript

## Acknowledgements

With thanks to David Bartle Paediatric Consultant Royal Devon University Healthcare NHS Foundation Trust for their advice on the protocol. Tanya Hynd and Isabelle Hawksworth from the PPIE team from NIHR ARC South West Peninsula for helping to recruit and organise the PPI group and members of the PPI group for their input throughout the review as described above.

## CRediT statement

Rebecca Whear: conceptualisation, methodology, analysis, investigation, writing original draft, reviewing and editing.

Becca Abbott: conceptualisation, methodology, analysis, investigation, writing original draft, reviewing and editing.

Harriet Aughey: conceptualisation, writing original draft, reviewing and editing.

Morwenna Rogers: conceptualisation, methodology, analysis, investigation, writing original draft, reviewing and editing.

Alison Bethel: conceptualisation, methodology, analysis, investigation, writing original draft, reviewing and editing.

Stuart Logan: conceptualisation, writing original draft, reviewing and editing. Kelly Boxall: conceptualisation, reviewing and editing.

Jo Thompson Coon: conceptualisation, methodology, analysis, investigation, writing original draft, reviewing and editing, supervision.

## Conflicts of Interest

Jo Thompson Coon, Rebecca Whear, Rebecca Abbott, Morwenna Rogers, Alison Bethel have no conflicts of interest to declare.

Harriet Aughey, Stuart Logan and Kelly Boxall were involved in a pilot study at the Royal Devon and Exeter (Wonford) Hospital (RD&E) site of the Royal Devon University Healthcare NHS Foundation Trust where a switch from IV to oral antibiotics after 36 hours of treatment for suspected EOS was piloted in 2024 and have contributed to the development of implementation resources available from Health Innovation South West.

### Box 1: Search strategy for Embase

Embase <1974 to 2025 April 23>

1. newborn/ 640530
2. newborn*.tw. 234040
3. (new adj2 born*).tw. 8056
4. neonat*.tw. 441845
5. (baby or babies).tw. 124145
6. 1 or 2 or 3 or 4 or 5 970749
7. oral drug administration/ 416217
8. 8 (oral* or buccal*).tw. 1224323
9. 9 ((enteral* or home or mouth or liquid*) adj3 (administrat* or therap* or treatment* or medication* or deliver* or dose* or dosage*)).tw. 42956
10. 7 or 8 or 9 1547534
11. Amox-clav*.tw. 59
12. amoxicillin*.tw. 32334
13. ampicillin*.tw. 36162
14. antibacteri*.tw. 152423
15. (anti adj2 bacteri*).tw. 10405
16. 16 antibiotic*.tw. 604955
17. (anti adj2 biotic*).tw. 460
18. antimicrob*.tw. 323288
19. augmentin*.tw. 29170
20. cefalexin*.tw. 696
21. cefpodoxim*.tw. 1499
22. chloramphenicol*.tw. 28708
23. cloxacillin*.tw. 2511
24. co amoxiclav*.tw. 1387
25. coamoxiclav*.tw. 211
26. flucloxacillin*.tw. 1888
27. nafcillin*.tw. 991
28. penicillin*.tw. 63943
29. antibiotic agent/ 428009
30. antibiotic therapy/ 171153
31. cefalexin/ 21739
32. chloramphenicol/ 72011
33. cloxacillin/ 11857
34. amoxicillin plus clavulanic acid/ 55577
35. flucloxacillin/ 10195
36. nafcillin/ 5671
37. penicillin derivative/ 56198
38. or/11-37 1293277
39. switch.ti. or (switch adj2 therap*).ab. 25579
40. 38 or 39 1318106
41. 6 and 10 and 40 2924
42. Animal experiment/ not (human experiment/ or human/) 2734909
43. 41 not 42 2747

---

**Supplementary Table 1:**
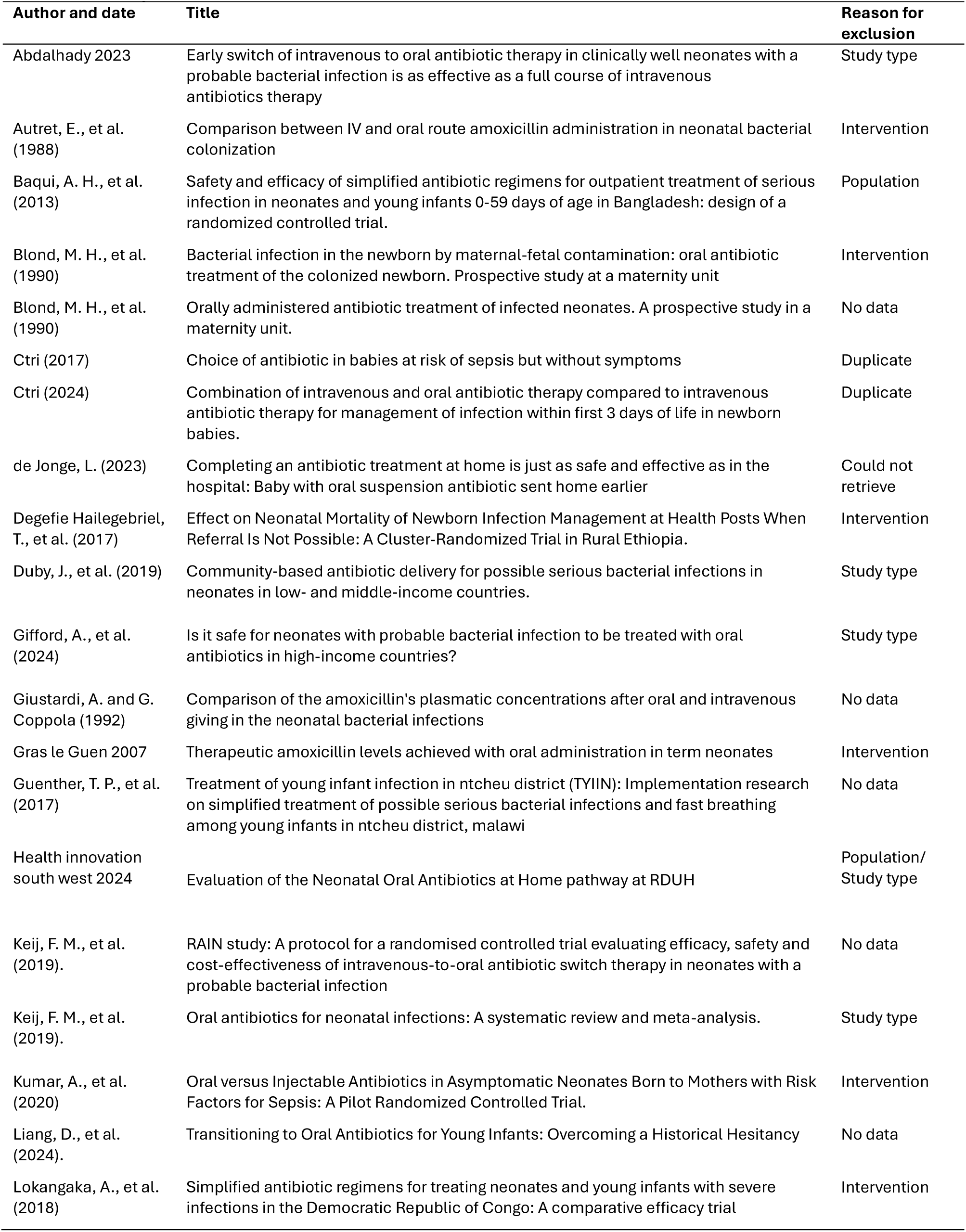
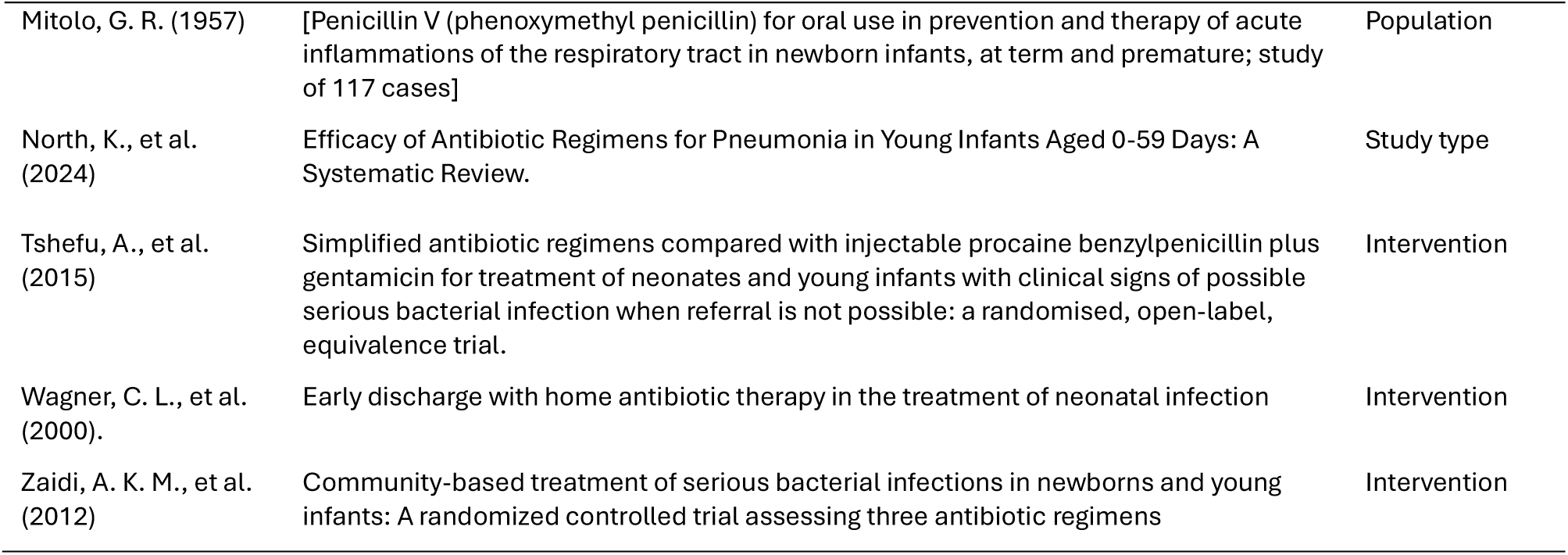
Excluded studies.

## References

1. Stark A, Smith PB, Hornik CP, et al. Medication Use in the Neonatal Intensive Care Unit and Changes from 2010 to 2018. J Pediatr 2022;240:66–71.e4. doi: 10.1016/j.jpeds.2021.08.075 [published Online First: 2021/09/06]

2. Jeferies AL. Management of term infants at increased risk for early-onset bacterial sepsis. Paediatrics & Child Health 2017;22(4):223–28. doi: 10.1093/pch/pxx023

3. Piyasena C, Galu S, Yoshida R, et al. Comparison of diagnoses of early-onset sepsis associated with use of Sepsis Risk Calculator versus NICE CG149: a prospective, population-wide cohort study in London, UK, 2020-2021. *BMJ Open* 2023;13(7):e072708. doi: 10.1136/bmjopen-2023-072708 [published Online First: 20230727]

4. National Institute for Health and Care Excellence. NICE guideline [NG195] Neonatal infection: antibiotics for prevention and treatment, 2021.

5. Giford A, Wooding EL, Ng KF. Is it safe for neonates with probable bacterial infection to be treated with oral antibiotics in high-income countries? Arch Dis Child 2024;109(8):681–87. doi: 10.1136/archdischild-2023-326496 [published Online First: 2024/02/03]

6. Keij FM, Kornelisse RF, Hartwig NG, et al. Oral antibiotics for neonatal infections: a systematic review and meta-analysis. J Antimicrob Chemother 2019;74(11):3150–61. doi: 10.1093/jac/dkz252 [published Online First: 2019/06/27]

7. McDermott JH, Wolf J, Hoshitsuki K, et al. Clinical Pharmacogenetics Implementation Consortium Guideline for the Use of Aminoglycosides Based on MT-RNR1 Genotype. Clinical Pharmacology & Therapeutics 2022;111(2):366–72. doi: 10.1002/cpt.2309

8. Page MJ, McKenzie JE, Bossuyt PM, et al. The PRISMA 2020 statement: an updated guideline for reporting systematic reviews. BMJ 2021;372:n71. doi: 10.1136/bmj.n71

9. Whear R, Abbott R, Thompson Coon J, et al. Efectiveness, cost efectiveness and experiences of switching from intravenous to oral antibiotics in neonates with probable early onset bacterial infection: a systematic review. PROSPERO 2025. https://www.crd.york.ac.uk/PROSPERO/view/CRD420251044158.

10. Thomas BH, Ciliska D, Dobbins M, Micucci S. A process for systematically reviewing the literature: providing the research evidence for public health nursing interventions. Worldviews Evid Based Nurs 2004;1(3):176–84. doi: 10.1111/j.1524-475X.2004.04006.x [published Online First: 2006/12/14]

11. Keij FM, Kornelisse RF, Hartwig NG, et al. Eficacy and safety of switching from intravenous to oral antibiotics (amoxicillin-clavulanic acid) versus a full course of intravenous antibiotics in neonates with probable bacterial infection (RAIN): a multicentre, randomised, open-label, non-inferiority trial. The lancet Child & adolescent health 2022;6(11):799–809. doi: 10.1016/S2352-4642(22)00245-0

12. Campbell M, McKenzie JE, Sowden A, et al. Synthesis without meta-analysis (SWiM) in systematic reviews: reporting guideline. BMJ 2020;368:l6890. doi: 10.1136/bmj.l6890

13. O’Neill J, Tabish H, Welch V, et al. Applying an equity lens to interventions: using PROGRESS ensures consideration of socially stratifying factors to illuminate inequities in health. Journal of Clinical Epidemiology 2014;67(1):56–64. doi: 10.1016/j.jclinepi.2013.08.005

14. Gyllensvärd J, Ingemansson F, Hentz E, et al. C-reactive protein-and clinical symptoms-guided strategy in term neonates with early-onset sepsis reduced antibiotic use and hospital stay: a quality improvement initiative. BMC Pediatr 2020;20(1):531. doi: 10.1186/s12887-020-02426-w [published Online First: 2020/11/22]

15. Malchau Carlsen EL, Dungu KHS, Lewis A, et al. Switch from intravenous-to-oral antibiotics in neonatal probable and proven early-onset infection: A prospective population-based real-life multicentre cohort study. Archives of Disease in Childhood: Fetal and Neonatal Edition 2024;109(1):34 EP - 40. doi: 10.1136/archdischild-2023-325386

16. Manzoni P, Esposito S, Gallo E, et al. Switch therapy in full-term neonates with presumed or proven bacterial infection. J Chemother 2009;21(1):68–73. doi: 10.1179/joc.2009.21.1.68 [published Online First: 2009/03/20]

17. National Institute for Health and Care Excellence. Neonatal infection: antibiotics for prevention and treatment [NICE Guideline No. 195], 2021.

18. UK Health Security Agency. National criteria for prompt intravenous-to-oral switch (IVOS) of antimicrobials in children and young people (including newborns): Gov.uk; 2024 [Available from: https://www.gov.uk/government/publications/antimicrobial-intravenous-to-oral-switch-criteria-for-early-switch/national-criteria-for-prompt-intravenous-to-oral-switch-ivos-of-antimicrobials-in-children-and-young-people-including-newborns2025.

